# Subconcussive head impact exposure differences between drill intensities in U.S. high school football

**DOI:** 10.1101/2020.03.29.20047167

**Authors:** Kyle Kercher, Jesse A. Steinfeldt, Jonathan T. Macy, Keisuke Ejima, Keisuke Kawata

## Abstract

**Purpose:** USA Football established five levels of contact (LOC) to guide the intensity of high school football practices. However, it remains unclear whether head impact exposure differs by LOC. The purpose of this study was to examine head impact frequency and magnitude by LOC in the overall sample and three position groups.

**Methods:** This longitudinal observational study included 24 high school football players during all practices and games in the 2019 season. Players wore a sensor-installed mouthguard that monitored head impact frequency, peak linear acceleration (PLA), and rotational head acceleration (PRA). Practice/game drills were filmed and categorized into 5 LOCs (*air, bags, control, thud, live*), and head impact data were assigned into 5 LOCs. Player position was categorized into linemen, hybrid, and skill.

**Results:** A total of 6016 head impacts were recorded during 5 LOCs throughout the season. In the overall sample, total number of impacts, sum of PLA, and PRA per player increased in an incremental manner (*air*<*bag*s<c*ontrol*<*thud*<*live*), with the most head impacts in *live* (113.7±17.8 hits/player) and the least head impacts in *air* (7.7±1.9 hits/player). The linemen and hybrid groups had consistently higher impact exposure than the skill group. Average head impact magnitudes by position group were higher during *live* drills (PLA (41.0-45.9*g*) and PRA (3.3-4.6 krad/s^2^) per head impact), whereas other LOCs had lower magnitudes (PLA (18.2-23.2*g*) and PRA (1.6-2.3krad/s^2^) per impact).

**Conclusion:** Our data suggest that LOC may influence cumulative head impact exposure in high school football, with players incurring frequent head impacts during *live, thud*, and *control*. The data indicate the importance of considering LOCs to refine practice guidelines and policies to minimize head impact burden in high school football athletes.

## INTRODUCTION

The long-term consequence of sport-related head injury is a complex public health issue with no concrete solution (1, 2). Despite inherent risk of head injury in contact sports (e.g., American football, hockey, soccer), participating in these team sports, especially during developmental age, provides well-documented benefits, including higher levels of physical activity, improved mental health, and lower likelihood of smoking cigarettes and using illegal substances (3). In an attempt to promote a safer environment, youth football organizations have implemented the teaching of proper tackling techniques and reduced the size of the field and number of players on the field (4). These modifications are further substantiated in high school, college, and professional football, where kickoffs and touchbacks were adjusted to reduce injuries (5), well-designed protective helmets are worn, and tackling of defenseless players and blindside blocking are prohibited (6, 7). Consistent with these adjustments, concussion and catastrophic injury rates have been reduced (8, 9); however, subconcussive head impact exposure has proven more complex.

Subconcussive head impact is defined as a hit to the head that does not induce overt concussion symptoms (10). These head impacts are most common in American football, where athletes can experience several hundred impacts with some exceeding 1,000 head impacts in a single season (11). Evidence has emerged to indicate that both high school and college football players with frequent subconcussive head impact exposure exhibit neuronal microstructural damage (12, 13), abnormal brain activation (14, 15), ocular-motor impairment (16), and elevation in brain-injury blood biomarkers (17). One line of research suggests that long-term exposure to these hits is a key factor in developing neurodegenerative disorders later in life (18, 19). Although USA Football (for high school) and the NCAA (for college) have eliminated practicing two times in the same day (two-a-days) to minimize head impact exposure, one study found that total head impact frequency during a summer camp increased by 26% (20). Similarly, a policy change to reduce the number of preseason practices from 29 to 25 failed to reduce head impact exposure in college football players, with one team’s cumulative head impacts increasing up to 35% (21). These mixed results highlight the need to dissect football practices to increase our understanding of what type of contact drills and intensities cause the greatest exposure to subconcussive head impacts.

USA Football (the national governing body over amateur football) has identified five levels of contact (LOCs) that define the intensities and structure of football practices nationwide. The LOCs were designed to guide effective practice schedules through a step-by-step approach to teach fundamental football skills (22). The National Federation of State High School Associations implemented the LOCs in their high school football practice guidelines beginning in 2014. However, it remains unknown whether, and to what extent, different levels of contact influence head impact exposure in high school football players across an entire season and between position groups.

Therefore, we conducted a longitudinal observational study to examine cumulative head impact exposure across different drill intensities in high school football players over the course of a single season. Our primary hypothesis was that there would be an incremental increase in cumulative head impact kinematics between LOCs in the overall sample with *live* recording the greatest head impact exposure and air recording the lowest: *air* < *bags* < *control* < *thud* < *live*. Since the proximity to opponents and nature of contact during nearly every play for linemen (23), we also hypothesized that linemen would have greater head impact exposure in most LOCs, compared to the hybrid and skill positions. Our secondary aim was to identify the average head impact magnitudes by LOC and position group. Lastly, we aimed to explore frequency of head impacts that were within 25-60 *g*, 60-100 *g*, or >100 *g* in each LOC.

## METHODS

### Participants

This single-site, observational study included 24 male high school football players at Bloomington High School-North. The study was conducted during the 2019 football season including practices and games during the pre-season, in-season, and playoffs. None of the 24 players was diagnosed with a concussion during the study period as confirmed by team athletic trainer and physician. Inclusion criterion was being an active football team member. Exclusion criteria included a history of head neck injury (including concussion) in the previous year or neurological disorders. The Indiana University Institutional Review Board and the Monroe County Community School Corporation Research Review Board approved the study, and all participants and their legal guardians gave written informed consent.

### Study Procedures

At the preseason data collection, self-reported demographic information (age, race/ethnicity, height, weight, number of previously diagnosed concussions, and years of experience in various contact sports, including tackle football) were obtained. Participants were custom-fitted with the Vector mouthguard (Athlete Intelligence, Inc.) that measured the number of hits and magnitude of head linear and rotational acceleration. Participants wore the mouthguard for all practices and games from pre-season training camp (August 13, 2019) to the end of the season (November 1, 2019). Head impact data were collected during 34 practices, 11 varsity games, and 9 junior varsity games over a 12-week season. Video data were collected using Hudl (Agile Sports Technologies, Inc.) during the same timeframe as subconcussive head impact data collection. Participants’ playing positions were verified by team coaches and categorized into three groups as follows: 11 linemen athletes (defensive lineman, offensive lineman), 7 hybrid athletes (tight end, linebacker, running back), and 6 skill athletes (wide receiver, defensive back), which is in line with prior literature (24, 25). No quarterbacks were included in this study. In accordance with USA Football guidelines (26), head impacts were categorized by LOC: *air, bags, control, thud*, and *live*. See Levels of Contact and Film Review section for more details and supplemental file A for example video for each LOC.

### Head Impact Measurement

This study used an instrumented Vector mouthguard for measuring frequency of head impacts as well as linear and rotational head kinematics during impacts, as previously described (27). The mouthguard employs a triaxial accelerometer (ADXL377, Analog Devices) with 200 *g* maximum per axis to sense linear acceleration. For rotational kinematics, a triaxial gyroscope (L3GD20H, ST Microelectrics) was employed. An impact is detected when a linear acceleration magnitude exceeds 10.0 *g* for three consecutive samples (sampling every 0.2 milliseconds). All impact data with a standard hit duration of 96 milliseconds were transmitted wirelessly through the antenna transmitter to the sideline antenna and computer, then stored on a secure internet database. The Vector mouthguard is installed with an in-mouth sensor to ensure that data acquisition occurs only when the mouthguard is securely fitted in one’s mouth. Linear acceleration data were transformed within the Athlete Intelligence software to the head’s center of gravity based on the 50^th^ percentile male. From raw impact data extracted from the server, the number of hits, peak linear acceleration, and peak rotational acceleration were used for descriptive analyses. Timestamps of all impacts were used to correspond with timeframes of each LOC.

### Levels of Contact and Film Review

The five LOCs are *air, bags, control, thud*, and *live* with *air* being estimated to have the lowest intensity and *live* being the highest (26). *Air* is defined as drills being run unopposed and without contact. *Bags* is defined as drills being run against a bag or soft-contact surface. *Control* is defined as drills being run at an assigned speed until the moment of contact. It does not involve tackling, rather contact is above the waist and players stay on their feet. *Thud* is defined as drills being run at a competitive, fast speed through the moment of contact. It does not involve full tackling, rather contact is above the waist and players stay on their feet and a quick whistle ends the drills. *Live* is defined as drills being run in game-like conditions that include live-drill during practice as well as real games. *Live* should be the only time players are allowed to fully tackle another player to the ground. All head impacts in *air, bags, control*, and *thud* were during practices, whereas *live* occurred in both practices and games.

Daily subconcussive head impact exposures were quantified in our full sample of 24 football players. Level of contact was assigned to each of the head impacts by corresponding the real-time, verified practice plan to the real-time head impact data set. Based on our preliminary analysis, there was a strong agreement on the head impact occurrence between the Vector mouthguard and film analysis.

### Statistical Analysis

A series of descriptive analyses was conducted to examine all aims. Our outcome variables for primary and secondary aims were head impact count, sum of peak linear acceleration, and sum of peak rotational acceleration. These outcomes were expressed as mean ± standard error of means and assessed by 5 LOCs (*air, bags, control, thud*, and *live*) and 3 position groups (lineman, hybrid, and skill). For the exploratory aim, a total number of head impact within ranges of 25-60 *g*, 60-100 *g*, or >100 *g* in each LOC was assessed in the overall sample.

## RESULTS

### Demographics and overall head impact exposure

A total of 6016 head impacts were recorded during 5 LOCs from 24 high school football players throughout the season, resulting in an average of 250.7 hits, 5891.0 *g*, and 528.5 krad/s^2^. These data are not reflective of head impacts that occurred outside the 5 LOCs, such as walk-through and pre-practice/game conditioning. Demographics and head impact data in the overall sample are detailed in Table 1.

**Table 1.**
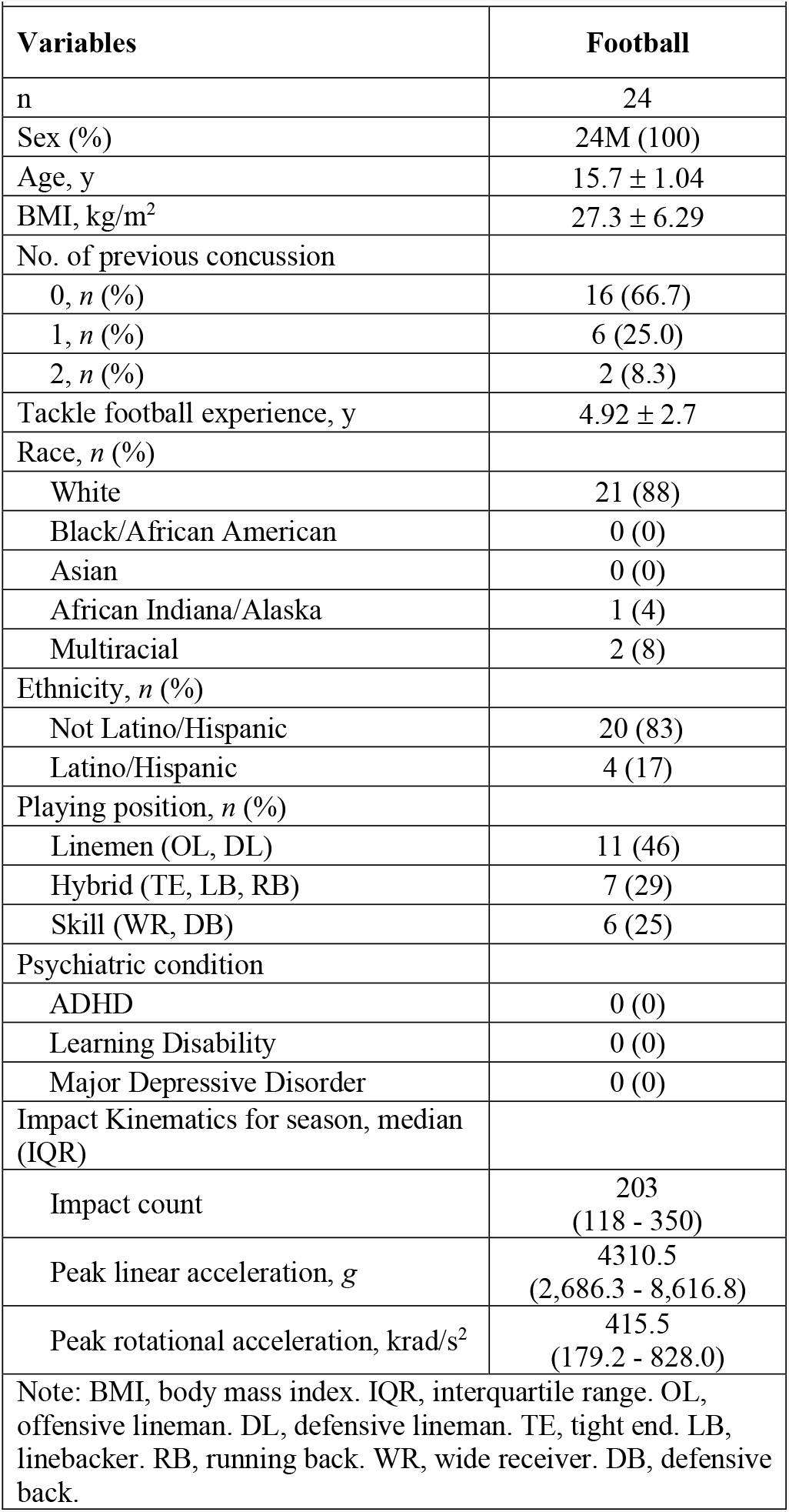
Group demographics and head impact kinematics.

### LOC-dependent cumulative head impact exposure

LOCs displayed an influence on head impact exposure in high school football players, as illustrated by the increasing cumulative exposure corresponding to increased levels of contact. Specifically, in the overall sample, total number of impacts per player as well as sum of PLA and PRA increased in an incremental manner (*air* < *bags* < *control* < *thud* < *live*), with the most head impacts being observed in live (113.7 ± 17.8 hits, 2,657.6 ± 432.0 *g*, 233.9 ± 40.1 krad/s^2^) and the least head impact exposure in air (7.7 ± 1.9 hits, 176.9 ± 42.5 *g*, 16.7 ± 4.2 krad/s^2^) (Figure 1A-C). Of note, *live* impacts were primarily during games (N=2,512, 92.08%) as opposed to during practices (N=216, 7.92%).

**Figure 1:**
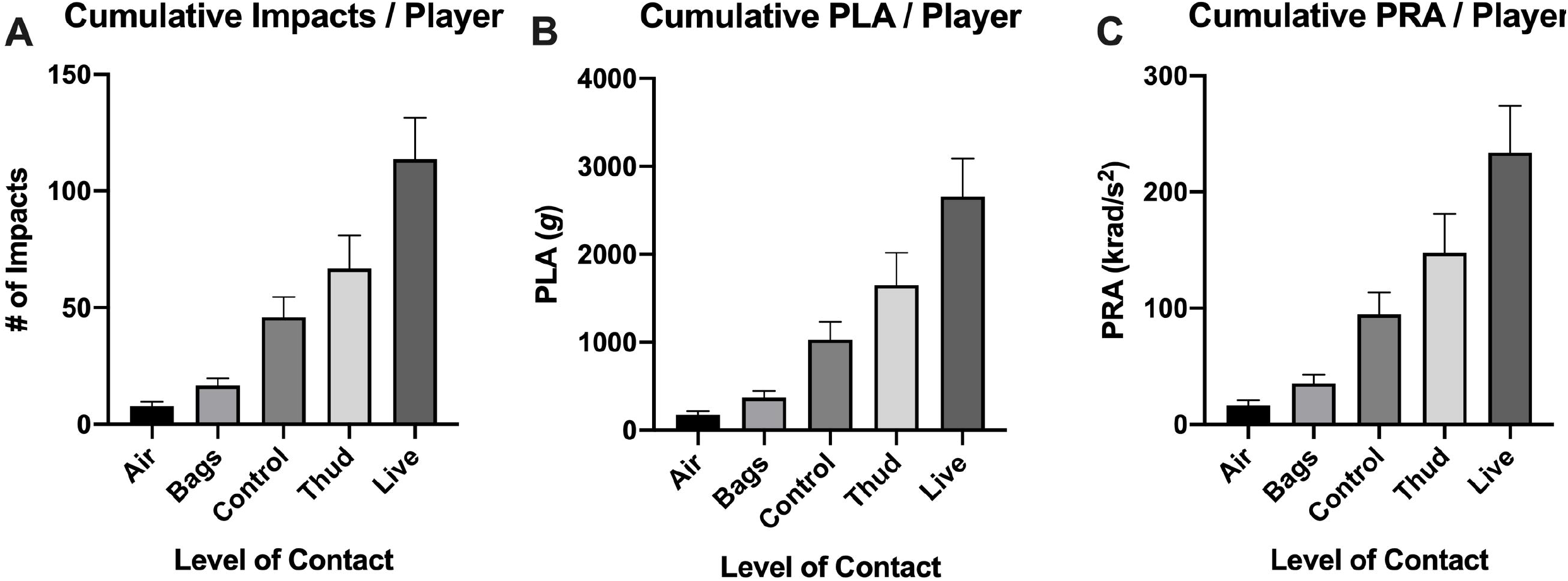
Cumulative head impact kinematics between LOC throughout a season. Cumulative (A) head impact count, (B) peak linear acceleration, and (C) peak rotational acceleration per player was influenced by LOC in an incremental manner, with *live* being the highest and *air* being the lowest.

All 3 position groups exhibited similar incremental patterns in head impact kinematics (*air* < *bags* < *control* < *thud* < *live*). However, the linemen and hybrid groups had consistently higher exposure in both frequency and magnitudes than the skill group across all LOCs (Figure 2A-C). Head impact kinematics for each LOC are detailed in Supplemental Table 1.

**Figure 2:**
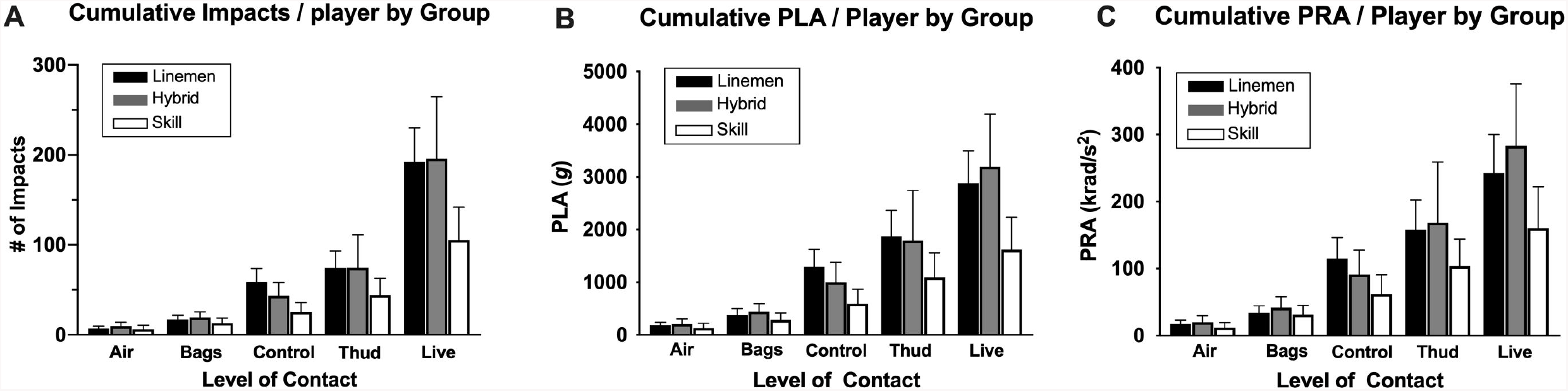
Group-dependent cumulative head impact exposure between LOC. All 3 groups shared similar incremental pattern in cumulative (A) head impact count, (B) peak linear acceleration, and (C) peak rotational acceleration per player, with *live* being the highest and *air* being the lowest. The linemen and hybrid groups exhibited consistently higher head impact exposure compared to the skill group.

### Head impact magnitudes by position group and LOC

Average head impact magnitudes by position group were shown to be similar across LOCs for *air, bags, control*, and *thud*, ranging between 18.2 and 23.2 g for PLA and 1.6 and 2.3 krad/s^2^ for PRA per head impact. However, *live* showed two times as high of PLA (41.0 to 45.9 *g*: Figure 3A) and PRA (3.3 to 4.6 krad/s^2^: Figure 3B) per head impact as other LOCs. The same pattern was observed in all 3 groups. Consistent with published papers (13, 23, 28), it was evident that there was a large number of head impacts across all LOCs that were within 10 to 30 *g*, which might have diluted the minority of high magnitude head impacts. Our exploratory analysis revealed that in the overall sample, high magnitude head impacts (60-100 *g* and >100 *g*) were most prevalent in *live*, followed by *thud, control, bags*, and then *air* (Figure 4A-C).

**Figure 3:**
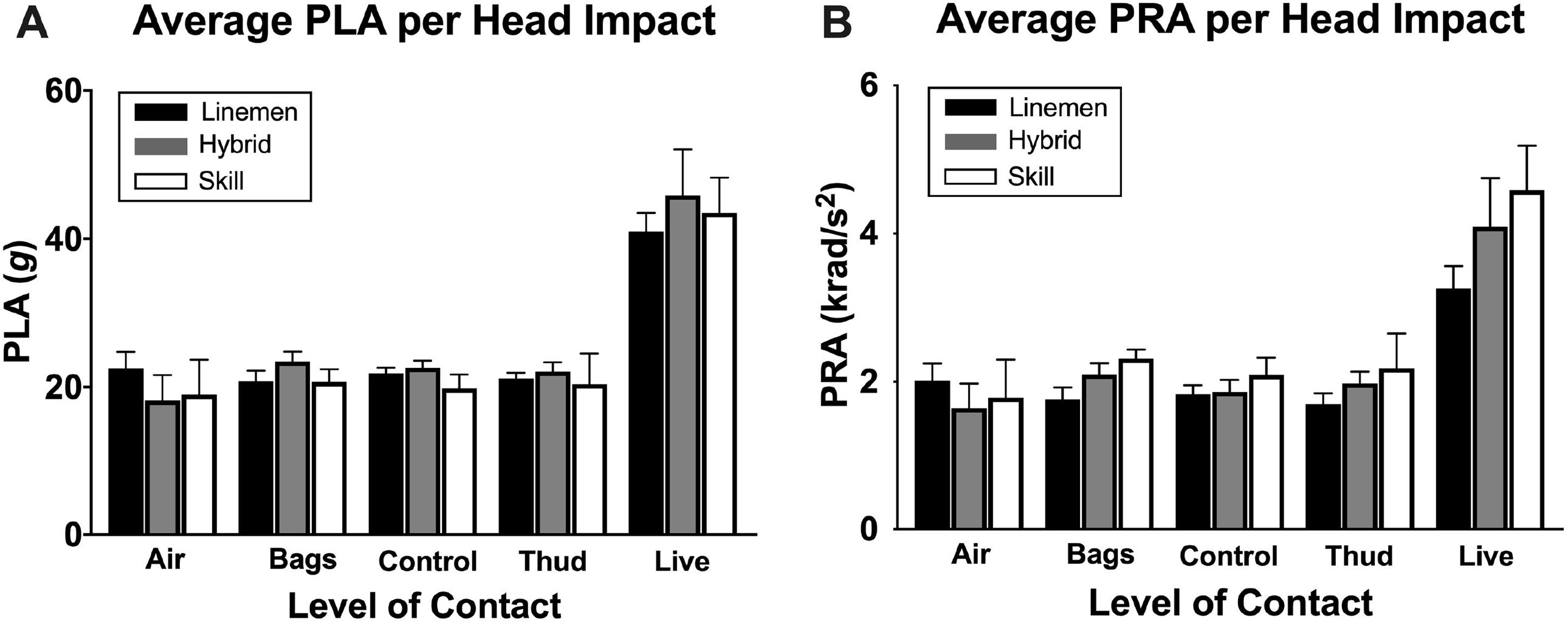
Average head impact magnitude per impact between LOC and group. Live showed nearly times as high head impact magnitude (A. peak linear acceleration; B. peak rotational acceleration) as 5 other LOC.

**Figure 4:**
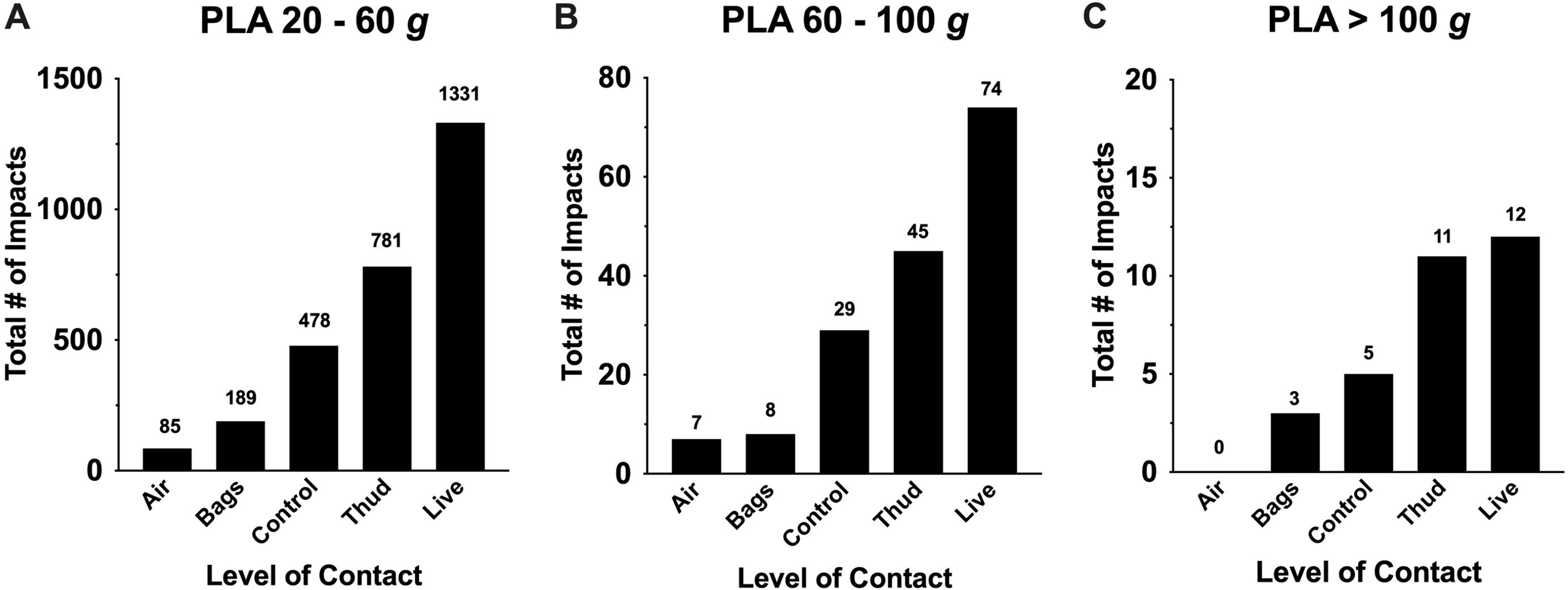
Frequency of head impacts within various magnitude range. A total number of head impacts from the overall sample throughout a season was categorized into peak linear acceleration ranging (A) 20 – 60 *g*, (B) 60-100 *g*, and (C) > 100 *g*. Live and thud consistently showed frequency head impacts in high impact magnitudes.

## DISCUSSION

Subconcussion research is still at its infancy, but it is a rapidly growing area of concern in sport injury prevention. To contribute to this emerging field, this study examined whether subconcussive head impact kinematics differed across LOCs and between player position groups. There were four key findings in this study. First, cumulative head impact exposure increased as the LOC increased, with frequent head impacts observed during *live, thud*, and *control* drills. Second, in line with literature (11), linemen and hybrid groups had consistently higher head impact exposure compared to the skill position group across all LOCs. Third, the mean head impact magnitude was similar (18 to 23 *g*) across *air, bags, control*, and *thud*, but head impacts during *live* showed greater magnitudes (40 to 45 *g*). Lastly, very high impact magnitudes (>100 *g*) were small in number overall but were more frequent in *thud* and *live* than other LOCs. Taken together, our data, for the first time, empirically support the USA football’s categorization of LOC while calling for a need to dissect football practice guidelines to make more specific recommendations for practice and games to minimize cumulative burden on adolescents’ brain health.

Owing to the sensor-installed helmets, mouthguards, headbands, and skin patches, our knowledge of head impact exposure in American football has drastically improved in the past 15 years. These technological advancements allowed researchers to evaluate head impact frequency and magnitude in various position groups, practice types (e.g., shell-only, full-gear), play types (e.g., running, passing, special teams), and time-based hit rates (29-34). Previous research has suggested that overall head impact exposure elevates in relation to increased practice duration, contact intensity, and time spent in high risk drills (21, 32). However, these variables differ greatly between players and position groups. For example, previous literature demonstrated that the differences between individual players accounted for 48% of the variance in head impact exposure during practice (35). Additionally, recent research suggests different types of plays (e.g., running, passing, special teams) also have different average head impact magnitudes which will influence cumulative head impact exposure (34). Despite this high degree of variance between players and play types, the linemen and hybrid position groups consistently have higher head impact exposure compared to skill positions such as receivers, defensive backs, and quarterbacks (30, 31). We were able to corroborate these observations in our study in the context of five different LOC, where frequent head impacts were notable especially during high intensity drills (*live, thud, control*), with linemen and hybrid players experiencing more head impacts than skill players.

Another major finding from this study was that *live* had the highest average PLA and PRA compared to *air, bags, control*, and *thud*, which shared similar impact magnitudes. Furthermore, strong magnitudes of head impacts (60 – 100 *g* and >100 *g*) were frequently observed during *live* and *thud*. These findings suggest current football guidelines that recommend limiting full contact (*live* & *thud*) during practice are appropriate. In fact, the football team in this study significantly minimized *live* drills during practice (n=216 impacts); therefore, 92% of head impacts in *live* was recorded during the games (n=2,512 impacts). In alignment with this idea, there have been attempts by various football governing bodies to restrict the amount of full contact in practices (20, 21, 26, 31, 36). For example, USA Football’s National Practice Guidelines for Youth Tackle Football suggest that full-contact should not be done for more than 30 minutes per day and no more than 90 minutes per week during the regular season (26). By limiting full contact practices from 3 days to 2 days per week, there was an average decline of 42% in head impact exposure in high school football players, albeit average magnitude of head impact remained unchanged (31). Some of these positive changes, however, do not translate into the college setting. Even though the NCAA eliminated two-a-day practices and reduced practice frequency from 29 to 25 practices during summer camp, to compensate for the loss of practice times, football teams tended to incorporate more high-intensity, contact-prone practice drills, leading to increased head impact exposure (20, 21). These data further substantiate the importance of regulating the duration of specific drill types, rather than restricting practice type and frequency.

There are several limitations to this study. Our examination of head impacts in high school football is limited in that it was conducted on a single high school football team in the Midwest composed of primarily white males; therefore, the results are not generalizable to the broader U.S. population of high school football teams. A second limitation of the study is that the USA Football LOC guidelines are just that, guidelines. They are not legislation, and it is unknown what percentage of high schools currently utilize the USA Football LOC system; thus, this further limits the generalizability. Implementation of the LOCs is not a perfectly reliable variable in that situations occur in practice LOCs that may overlap with other LOCs. Coaches in this high school attempted to implement the LOCs consistently, but to control up to 22 athletes participating in a single drill at the same time is not always feasible. These limitations portend a follow-up research question as to whether regulating the number of repetitions in each LOC is a more effective approach than teamwide contact restrictions for minimizing the frequency and magnitude of subconcussive head impacts in individual players (or position groups). Hence, for future research we suggest examining subject-specific head impact data that incorporate multiple predictor variables such as the repetitions per player per drill, LOC, position group, and impacts per event (i.e., drill, practice/game, or week) for head impact kinematic outcomes.

## CONCLUSION

Our data suggest that the LOC may influence cumulative head impact exposure in high school football players, with players incurring frequent head impacts during *live, thud*, and *control*. Strong magnitudes of head impacts were frequently observed during *live* and *thud*. Many head impact investigations and contact recommendations have focused on limiting the number of practices (20, 21, 37), time involved in full contact in general (26, 30, 38), or changing game rules (8). Our data indicate the importance of considering LOCs to refine football practice guidelines/policies to minimize cumulative head impact burden in high school football players.

## Data Availability

Data can be released up on contacting the corresponding author.

